# Development and multicenter external validation of A Data-Driven Scoring System for Early and Rapid Identification of Sepsis in Emergency Departments

**DOI:** 10.1101/2025.09.27.25336784

**Authors:** Yanwei Jin, Yinzhao Wang, Xiaodong Huang, David A. Wacker, Michael A. Puskarich, Feng Xie

## Abstract

**Importance:** Sepsis is a leading cause of morbidity and mortality worldwide. Timely recognition and treatment in the emergency department (ED), often referred to as the “golden window,” are critical to improving outcomes. Yet, current diagnostic tools either demonstrate limited accuracy or rely on laboratory results that are not immediately available during initial ED evaluation, constraining rapid and reliable sepsis identification in the ED.

**Objective:** To develop and externally validate a data-driven interpretable score for early identification of sepsis in the ED across three large health systems.

**Design, Setting, and Participants:** This retrospective cohort study used electronic health records from three health systems. The primary derivation cohort included all ED visits at 11 hospitals within the M Health Fairview system (Minnesota, 2019-2025). Two external cohorts include ED visits from the Beth Israel Deaconess Medical Center (BIDMC, Boston, 2011-2019) extracted from the MIMIC-IV-ED database, and ED visits from the Stanford Health Care (Stanford, 2020-2022) sourced from the MC-MED database. In our analysis, completed in August 2025, we developed the Emergency Sepsis Risk Prediction (ESRP) score using the AutoScore framework. We evaluated its performance against commonly used bedside tools, including quick Sequential Organ Failure Assessment (qSOFA), National Early Warning Score (NEWS), the Modified Early Warning Score (MEWS), and the Rapid Emergency Medicine Score (REMS), as well as logistic regression (LR) and random forest (RF) models.

**Main Outcomes and Measures:** The primary outcome was sepsis diagnosis during the ED or hospital stay, determined from ICD-9 and ICD-10 discharge codes. Model performance was evaluated using the area under the receiver operating characteristic curve (AUROC) and the area under the precision–recall curve (AUPRC), sensitivity, specificity, positive predictive value (PPV), and negative predictive value (NPV).

**Results:** The study included a total of 2,193,244 ED visits across three sites: 1,626,055 in the Minnesota cohort (1,271,865 for model derivation, and 354,190 for internal validation), and 448,804 and 118,385 in the BIDMC and Stanford external validation cohorts, respectively. In Minnesota internal validation, the ESRP score achieved an AUROC of 0.820 (95% CI, 0.810– 0.825) and an AUPRC of 0.054 (95% CI, 0.051–0.058). In the BIDMC cohort, the ESRP achieved an AUROC of 0.838 (95% CI, 0.833–0.842), compared with 0.636 (95% CI, 0.633– 0.640) for qSOFA and 0.760 (95% CI, 0.757–0.765) for NEWS. In the Stanford cohort, the ESRP achieved an AUROC of 0.892 (95% CI, 0.887–0.898), compared with 0.697 (95% CI, 0.684–0.716) for qSOFA and 0.870 (95% CI, 0.861–0.881) for NEWS.

**Conclusions:** The ESRP score, based on 10 easily obtainable triage variables, provided accurate, generalizable, and interpretable early sepsis identification across diverse ED populations. Its simplicity and strong performance suggest potential for integration into routine ED triage workflows to support timely sepsis care.

## Introduction

Sepsis, a life-threatening organ dysfunction caused by a dysregulated host response to infection, represents a critical challenge in emergency medicine. The emergency department (ED) is the first point of contact for up to 80% of sepsis patients, with 86% of all hospitalized cases being identified upon admission^1^. This positions the ED as the primary battleground for early detection and intervention. The high-pressure ED environment, characterized by high patient volume and rapid clinical decision-making, demands the timely recognition of sepsis to improve patient outcomes and reduce mortality. The consequences of delayed diagnosis are severe, with mortality risk increasing by approximately 7.6% for each hour that effective treatment is delayed^2^. Consequently, the earliest possible identification of at-risk patients represents the most critical step in the care pathway. This initial assessment dictates crucial subsequent decisions, including the initiation of time-sensitive sepsis cares and hospital admission. This process is invariably complicated by the necessity of making high-stakes decisions with incomplete clinical data, as clinicians cannot afford to wait for comprehensive laboratory results. The diagnostic challenge is further exacerbated by the heterogeneous nature of sepsis^3^. It can arise from diverse etiologies, such as community-acquired pneumonia or healthcare-associated infections, and often manifests in vulnerable populations, including the elderly and immunocompromised. Furthermore, the clinical presentation is frequently nonspecific, with subtle signs like general fatigue or altered mental status, which can obscure an accurate and swift diagnosis.

Existing prediction tools are inadequately suited for the unique demands of the ED. Scores such as the Sequential Organ Failure Assessment (SOFA) are designed for inpatient and ICU settings and rely on laboratory values that are not immediately available at ED triage^4^. The simplified quick SOFA (qSOFA) score, while faster, suffers from low diagnostic accuracy^5^. Other risk stratification systems like the National Early Warning Score (NEWS) and the Rapid Emergency Medicine Score (REMS) were designed for ED, but not specifically for sepsis risk assessment^6,7^. Although sophisticated machine learning models can achieve higher accuracy, their practical application in the ED is challenging because complex models like Random Forest (RF), Gradient Boosting (GB), and Multilayer Perceptrons (MLP) are often considered “black-box” models due to their lack of interpretability^8^. This opacity, along with potentially high computational demands, can limit their suitability for the fast-paced ED environment and risk increasing the burden on clinicians^9^, especially for low-resource settings To address this gap, we developed and validated the Emergency Sepsis Risk Prediction (ESRP) scoring system. The ESRP is a novel, parsimonious tool that leverages ten readily available variables obtainable at triage to provide an immediate risk assessment. Designed for seamless integration into existing ED workflows without imposing an additional burden, the ESRP empowers clinicians with a rapid, accurate, and actionable tool to guide decision-making, optimize patient care, and ultimately improve outcomes for patients with suspected sepsis.

## Methods

### Study Design and Data Sources

This retrospective cohort study utilized electronic health record (EHR) data from three sites. The primary derivation cohort included all ED visits within the M Health Fairview system under University of Minnesota (UMN), between January 1, 2019, and January 22, 2025. This dataset encompasses patient visits across 11 distinct EDs in Minnesota, representing a diverse patient mix from urban, suburban, and rural settings. External validation was performed using two additional datasets: (1) the Medical Information Mart for Intensive Care IV–Emergency Department (MIMIC-IV-ED)^10,11^ dataset, containing over 450,000 ED visits from the Beth Israel Deaconess Medical Center (BIDMC) in Boston between 2011 and 2019; (2) Multimodal Clinical Monitoring in the Emergency Department (MC-MED)^12,13^ dataset, comprising 118,385 adult ED visits from the Stanford Health Care between September 2020 and September 2022. This study was approved by the Institutional Review Board of the University of Minnesota (IRB #STUDY00023702).

### Cohort Selection and Data Preprocessing

We included adult patients aged 18 years or older. Visits with missing data on patient demographic, ED arrival information, or location were excluded. Our data preprocessing pipeline began with structuring the raw data extracted for each visit—including demographics, event timestamps, historical visit information, triage vitals, acuity scores, chief complaints, diagnoses, and medication history. From this, key predictive features were engineered, such as calculating the frequency of ED visits within a specific period and the time interval between consecutive visits. Missing vital signs were imputed using median values, with details provided in eMethods. Pain scores were imputed as 0, and Glasgow Coma Scale (GCS) scores were imputed as 15^14,15^. Comorbidity histories were consolidated by calculating the Charlson Comorbidity Index (CCI) and Elixhauser Comorbidity Index (ECI) based on all International Classification of Diseases (ICD)-9-CM and ICD-10-CM codes from the preceding five years. The prevalence of individual comorbidities captured by these indices ranged from approximately 1% to 10% within the study cohort. This process resulted in a final set of 64 variables for model development, which are listed in eTable1.

### Outcome Definition

The primary outcome was sepsis diagnosis, identified by the presence of ICD-9/10-CM discharge codes (detailed in eTable 1) or a structured sepsis diagnosis field in the medical record. In the UMN cohort, diagnoses were captured if documented during the index ED visit or within the subsequent seven days. For the BIDMC and Stanford cohorts, we used the associated hospital admission diagnosis record. Visits not meeting these criteria were classified as non-sepsis.

### Model Development and Performance Evaluation

The model development dataset was constructed using UMN data from 2019 to 2023 for training and 2024 to 2025 for internal validation. The BIDMC and Stanford cohorts were held out entirely for external validation. We developed the ESRP score using AutoScore, a ML algorithm for generating interpretable clinical scores^16–18^. For benchmarking, the ESRP score was compared against Linear Regression (LR) and RF models. These models were trained on two feature sets: the full set of 64 variables, and the parsimonious set of 10 variables used in the ESRP score. Additionally, several established clinical risk scores were calculated for comparison: qSOFA, NEWS, REMS, and the Modified Early Warning Score (MEWS)^19^. Model performance was evaluated using the area under the receiver operating characteristic curve (AUROC), the area under the precision-recall curve (AUPRC), sensitivity (recall), specificity, positive predictive value (PPV), and negative predictive value (NPV)^20^. All metrics were reported with 95% confidence intervals (CIs), estimated using 200 bootstrap resamples. To assess the model’s robustness and generalizability, pre-specified subgroup analyses were performed on the internal validation set, with performance stratified by ED location (urban, suburban, and rural, see eTable2 for the list of hospitals). The number of variables used in each final model was also documented as a measure of model complexity.

## Results

### Cohort Characteristics

A total of 1,679,022 ED visits from the UMN dataset (2019–2025) were included in the final analysis, containing 13,446 sepsis positive cases (overall prevalence: 0.8%). The data were chronologically split into a training set (n = 1,324,832; sepsis positive rate: 0.8%) and a testing set (n = 354,190; sepsis positive rate: 0.8%). The testing set served as an internal validation cohort to evaluate model performance. For a detailed performance analysis, the testing set was further stratified by geographic location into urban (n = 55,705; sepsis positive rate: 0.7%), suburban (n = 200,000; sepsis positive rate: 0.9%), and rural (n = 93,169; sepsis positive rate: 0.8%) subgroups. For external validation, we included two distinct cohorts: the BIDMC dataset (2011–2019), comprising 448,804 visits with a significantly higher sepsis prevalence of 2.5% (n=11,066), and the Stanford dataset (2020–2022), with 118,385 visits and a sepsis prevalence of 0.7% (n=843). An overview of the study framework is provided in Figure 1.

**Figure 1.**
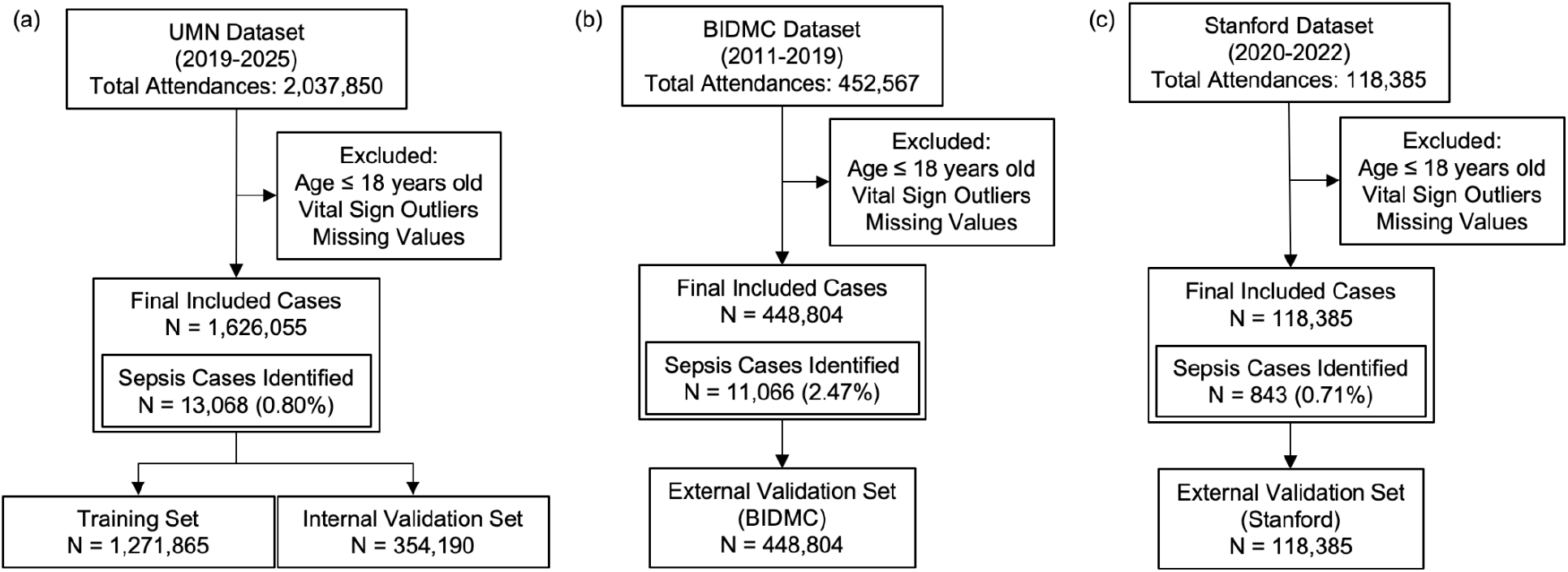
Flow of patient inclusion and cohort construction.

Demographic and clinical characteristics of the training, internal validation, and external validation cohorts are detailed in Table 1. The training and internal validation sets demonstrated similar demographic and clinical profiles, but a critical difference was observed in patient acuity as measured by the Emergency Severity Index (ESI). The BIDMC validation cohort presented with a significantly higher proportion of high-acuity patients; the combined percentage of patients triaged as ESI level 1 or 2 was 40.1% in the BIDMC validation cohort, nearly double the 23.2% seen in the internal validation cohort. Higher patient acuity is strongly correlated with a higher incidence of sepsis^21^. Notably, the sepsis prevalence in the BIDMC validation set was also significantly higher at 2.5%, compared to 0.8% and 0.7% in other cohorts, establishing a robust challenge for testing the model’s generalizability. Within the internal validation cohort, there were also observable differences among the geographic subgroups (detailed in eTable 3). These variations underscore the diverse patient populations represented in the dataset.

**Table 1.** Basic characteristics of the study cohort.

| Cohort | UMN -<br>Training set | UMN -<br>Testing set | BIDMC -<br>External<br>Validation | Stanford-External<br>Validation |
| --- | --- | --- | --- | --- |
| Sample Size (N) | 1,324,832 | 354,190 | 448,804 | 118,385 |
| Age, Mean (SD) (years) | 50.94 (20.85) | 52.07 (21.08) | 52.82 (20.62) | 53.26 (20.41) |
| Gender, Female (%) | 739,684 (55.8) | 199861 (56.4) | 242844 (54.1) | 64272 (54.3) |
| Gender, Male (%) | 585,148 (44.2) | 154329 (43.6) | 205960 (45.9) | 54077 (45.7) |
| ESI, Level 1 (%) | 8,492 (0.6) | 2269 (0.6) | 25363 (5.7) | 1066 (0.9) |
| ESI, Level 2 (%) | 262,466 (19.8) | 79936 (22.6) | 154545 (34.4) | 27880 (23.6) |
| ESI, Level 3 (%) | 797,940 (60.2) | 217173 (61.3) | 237565 (52.9) | 77786 (65.7) |
| ESI, Level 4 (%) | 236,538 (17.9) | 51402 (14.5) | 30160 (6.7) | 10973 (9.3) |
| ESI, Level 5 (%) | 19,396 (1.5) | 3410 (1.0) | 1171 (0.3) | 680 (0.6) |
| Systolic BP at Triage, Mean (SD) (mmHg) | 132.49 (20.30) | 132.60 (20.68) | 134.89 (21.96) | 135.79 (22.33) |
| Diastolic BP at Triage, Mean (SD) (mmHg) | 78.57 (13.93) | 78.22 (13.66) | 77.47 (14.59) | 79.06 (15.04) |
| Temperature at Triage, Mean (SD) (°C) | 36.70 (0.48) | 36.71 (0.43) | 36.71 (0.53) | 36.71 (0.45) |
| Heart Rate at Triage, Mean (SD) (bpm) | 80.34 (15.12) | 80.20 (15.24) | 85.08 (17.32) | 86.58 (18.39) |
| Oxygen Saturation at Triage, Mean (SD) (%) | 97.19 (2.98) | 97.17 (2.80) | 98.38 (2.40) | 98.31 (2.31) |
| Respiratory Rate at Triage, Mean (SD) (breaths/min) | 17.92 (3.64) | 18.21 (3.08) | 17.55 (2.47) | 18.20 (2.91) |
| Sepsis, Positive (%) | 10,477 (0.8) | 2969 (0.8) | 11066 (2.5) | 843 (0.71) |

**Abbreviations:** UMN, University of Minnesota; BIDMC, Beth Israel Deaconess Medical Center; ESI, Emergency Severity Index; BP, blood pressure; SD, standard deviation; bpm, beats per minute; mmHg, millimeters of mercury; °C, degrees Celsius. Sepsis, Positive (%) refers to the proportion of patients with sepsis diagnosed based on ICD-9/10 discharge codes.

### Development and Characteristics of the ESRP Score

We developed the ESRP score, a parsimonious clinical tool detailed in Table 2, which is calculated from ten readily available variables: age, a chief complaint of fever or chills, five initial triage vital signs (Heart Rate [HR], Systolic Blood Pressure [SBP], Diastolic Blood Pressure [DBP], Oxygen Saturation [SpO2], and Respiratory Rate [RR]), hospitalization within the past 90 days, and two specific Elixhauser comorbidities (Weight Loss and Uncomplicated Hypertension). The scoring system assigns the highest weights to age and presenting complaints: patients aged 50 or older receive 20 points, while a chief complaint of fever or chills contributes 19 points. Key vital sign abnormalities are also heavily weighted, significant hypotension (SBP < 90 mmHg) adds 13 points, and significant tachycardia (HR >= 110 bpm) adds 11 points. This design highlights that effective risk stratification for sepsis relies on a combination of baseline demographics (Age), presenting symptoms (Fever), and critical physiological derangements captured in the initial vital signs.

**Table 2.**
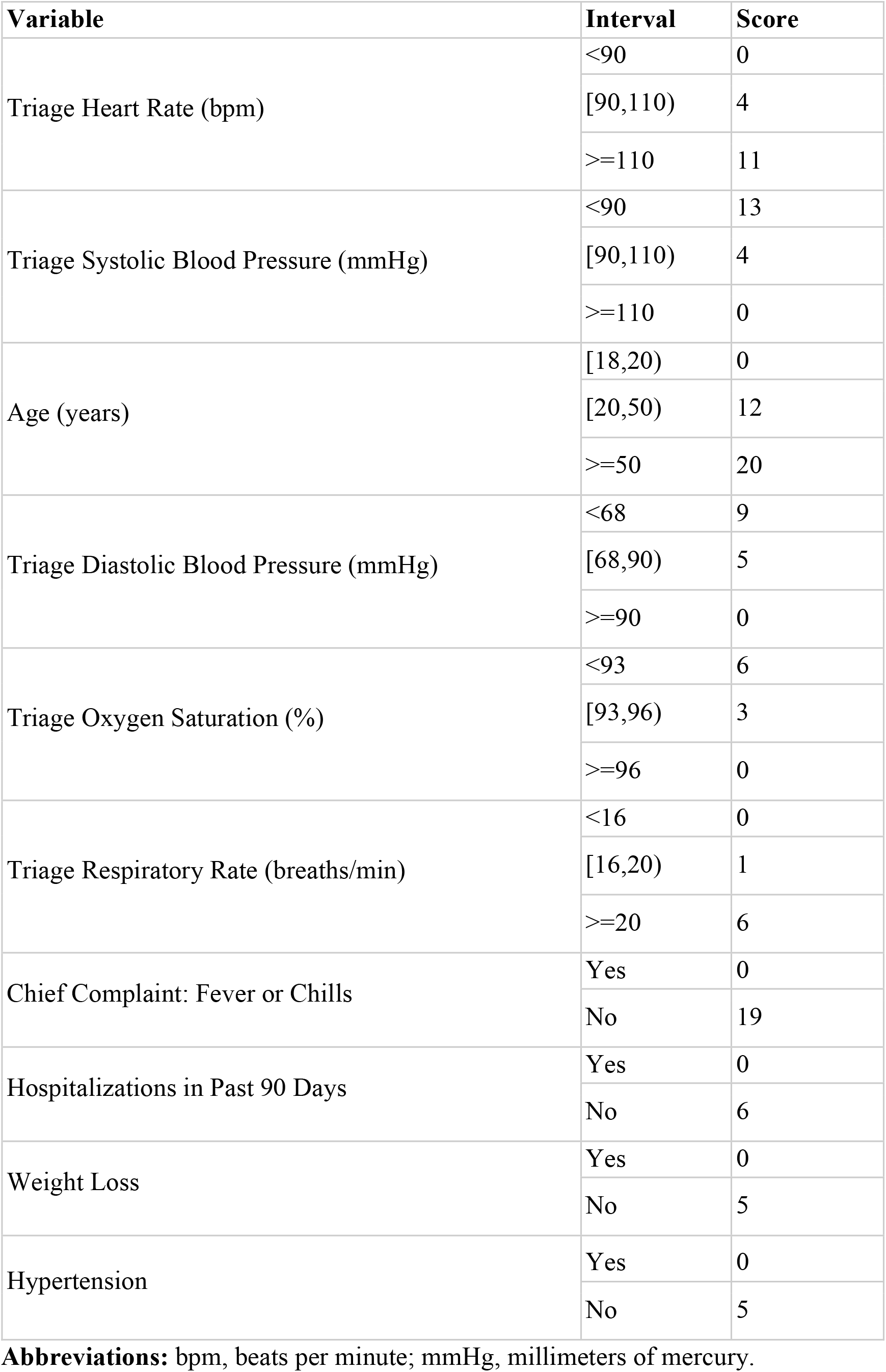
Score for Emergency Sepsis Risk Prediction (ESRP) Notes: Intervals are mutually exclusive and exhaustive. Chief Complaint refers to documentation at triage. Hospitalizations in the past 90 days include inpatient admissions only (excluding observation status stays). Weight loss and uncomplicated hypertension were defined based on past medical history.

### Model Performance

The predictive performance of the ESRP score was rigorously evaluated and compared against benchmark models and existing risk scores, with results summarized in Table 3 and Figure 2. Calibration analysis further demonstrated good agreement between predicted and observed risks after Platt scaling (eFigure1). On the internal validation cohort, the ESRP score demonstrated high overall discriminative ability, achieving an AUROC of 0.820 (95% CI: 0.810–0.825). However, recognizing that AUROC can be misleading in low-prevalence settings such as ours (~0.8%), we also assessed the AUPRC to provide a more complete performance evaluation^22^. The ESRP score achieved an AUPRC of 0.054 (95% CI, 0.051–0.058), substantially outperforming the prevalence baseline. At its optimal threshold, the score yielded a balanced sensitivity of 0.743 (95% CI, 0.732–0.786), indicating that approximately three-quarters of true sepsis cases were correctly identified (≈25% missed), and a specificity of 0.745 (95% CI, 0.705– 0.746), reflecting that roughly one-quarter of non-sepsis patients were incorrectly flagged. The PPV was 2.4%, consistent with the low underlying prevalence and highlighting a high rate of false alarms. In contrast, the NPV reached 99.7%, suggesting that patients predicted negative could be reliably ruled out as not having sepsis. This performance was comparable to the best-performing but more complex 64-variable LR model (AUROC 0.834, 95% CI: 0.830–0.839) and was decisively superior to other existing risk scores (AUROC range: 0.602–0.713). The ESRP model performed with high consistency in urban, suburban, and rural settings (detailed in eTable 4), achieving AUROCs of 0.805 (95% CI: 0.778–0.820), 0.815 (95% CI: 0.807–0.823), and 0.837 (95% CI: 0.829–0.851), respectively, indicating that its predictive ability is robust to the clinical setting. The ESRP score demonstrated strong generalizability, significantly outperforming all other evaluated clinical scores across two independent external validation cohorts. In the BIDMC cohort, the ESRP score achieved an AUROC of 0.838 (95% CI: 0.833– 0.842) and an AUPRC of 0.180 (95% CI: 0.173–0.186), significantly surpassing all other evaluated clinical scores. In the Stanford cohort, the ESRP score achieved an AUROC of 0.892 (95% CI: 0.887-0.898) and an AUPRC of 0.087 (95% CI: 0.068-0.101), significantly surpassing all other evaluated clinical scores.

**Table 3.** Comparison of performance for different models on the validation cohort.

| **UMN data - Internal Validation** |  |  |  |  |  |  |
| --- | --- | --- | --- | --- | --- | --- |
| **Model** | **AUROC** | **AUPRC** | **Threshold** | **Sensitivity** | **Specificity** | **Variables (N)** |
| LR - 10 Variables | 0.818 (0.813–0.823) | 0.053 (0.05–0.056) | 0.008 | 0.731 (0.726–0.782) | 0.759 (0.716–0.761) | 10 |
| RF - 10 Variables | 0.733 (0.723–0.741) | 0.038 (0.034–0.042) | 0.01 | 0.666 (0.651–0.678) | 0.739 (0.738–0.740) | 10 |
| LR - 64 Variables | 0.834 (0.83–0.839) | 0.052 (0.049–0.056) | 0.007 | 0.768 (0.751–0.777) | 0.757 (0.741–0.773) | 64 |
| RF - 64 Variables | 0.79 (0.782–0.796) | 0.065 (0.063–0.071) | 0.01 | 0.768 (0.751–0.778) | 0.713 (0.712–0.715) | 64 |
| ESI | 0.679 (0.673–0.686) | 0.015 (0.015–0.016) | 2 | 0.525 (0.513–0.539) | 0.77 (0.769–0.771) | 1 |
| qSOFA Score | 0.602 (0.598–0.61) | 0.016 (0.015–0.017) | 1 | 0.312 (0.305–0.327) | 0.888 (0.887–0.888) | 3 |
| ESRP Score | 0.82 (0.81–0.825) | 0.054 (0.051–0.058) | 36 | 0.743 (0.732–0.786) | 0.745 (0.705–0.746) | 10 |
| NEWS Score | 0.673 (0.665–0.683) | 0.022 (0.021–0.024) | 2 | 0.502 (0.489–0.516) | 0.77 (0.768–0.772) | 5 |
| REMS Score | 0.674 (0.666–0.684) | 0.015 (0.014–0.016) | 5 | 0.681 (0.666–0.696) | 0.606 (0.605–0.607) | 5 |
| MEWS Score | 0.603 (0.597–0.609) | 0.017 (0.016–0.019) | 2 | 0.383 (0.373–0.393) | 0.808 (0.806–0.809) | 4 |
| **BIDMC data - External Validation** |  |  |  |  |  |  |
| ESI | 0.721 (0.716–0.725) | 0.057 (0.056–0.058) | 2 | 0.747 (0.736–0.753) | 0.618 (0.617–0.619) | 1 |
| qSOFA Score | 0.636 (0.633–0.64) | 0.057 (0.055–0.059) | 1 | 0.345 (0.339–0.352) | 0.926 (0.925–0.926) | 2 |
| ESRP Score | 0.838 (0.833–0.842) | 0.18 (0.173–0.186) | 35 | 0.768 (0.758–0.776) | 0.752 (0.751–0.753) | 10 |
| NEWS Score | 0.76 (0.757–0.765) | 0.117 (0.113–0.122) | 2 | 0.648 (0.642–0.656) | 0.789 (0.788–0.79) | 5 |
| REMS Score | 0.706 (0.703-0.709) | 0.048 (0.047-0.05) | 5 | 0.719 (0.71-0.727) | 0.603 (0.602-0.605) | 5 |
| MEWS Score | 0.718 (0.713-0.723) | 0.093 (0.09-0.096) | 2 | 0.619 (0.612-0.628) | 0.768 (0.767-0.769) | 4 |
| **Stanford data - External Validation** |  |  |  |  |  |  |
| ESI | 0.75 (0.74-0.759) | 0.018 (0.016-0.019) | 2 | 0.709 (0.693-0.731) | 0.759 (0.756-0.761) | 1 |
| qSOFA Score | 0.697 (0.684-0.716) | 0.025 (0.021-0.03) | 1 | 0.503 (0.476-0.539) | 0.885 (0.884-0.886) | 2 |
| ESRP Score | 0.892 (0.887-0.898) | 0.087 (0.068-0.101) | 32 | 0.846 (0.795-0.868) | 0.792 (0.791-0.837) | 10 |
| NEWS Score | 0.87 (0.861-0.881) | 0.057 (0.048-0.064) | 2 | 0.865 (0.842-0.881) | 0.752 (0.75-0.754) | 5 |
| REMS Score | 0.746 (0.734-0.756) | 0.02 (0.018-0.024) | 6 | 0.632 (0.617-0.779) | 0.717 (0.581-0.719) | 5 |
| MEWS Score | 0.832 (0.819-0.849) | 0.061 (0.052-0.068) | 2 | 0.842 (0.768-0.866) | 0.715 (0.714-0.794) | 4 |

**Abbreviations:** LR = Logistic Regression; RF = Random Forest; AUROC = Area Under the Receiver Operating Characteristic Curve; AUPRC = Area Under the Precision–Recall Curve; ESI = Emergency Severity Index; qSOFA = Quick Sequential Organ Failure Assessment; ESRP = Emergency Sepsis Risk Prediction; NEWS = National Early Warning Score; REMS = Rapid Emergency Medicine Score; MEWS = Modified Early Warning Score; BIDMC = Beth Israel Deaconess Medical Center; UMN = University of Minnesota.

**Figure 2.**
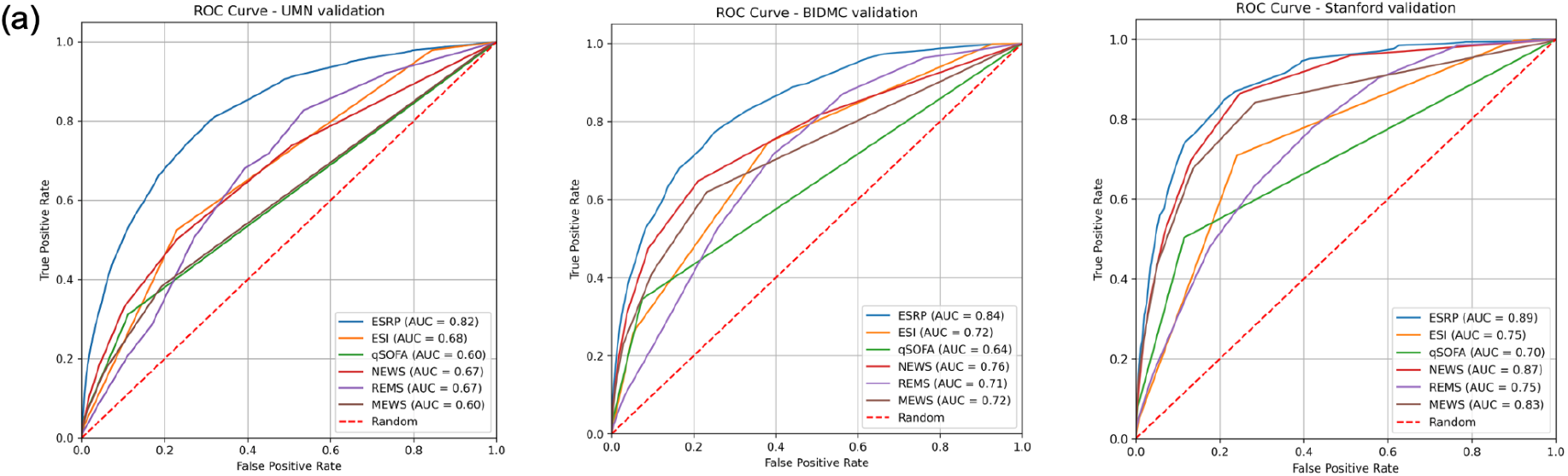
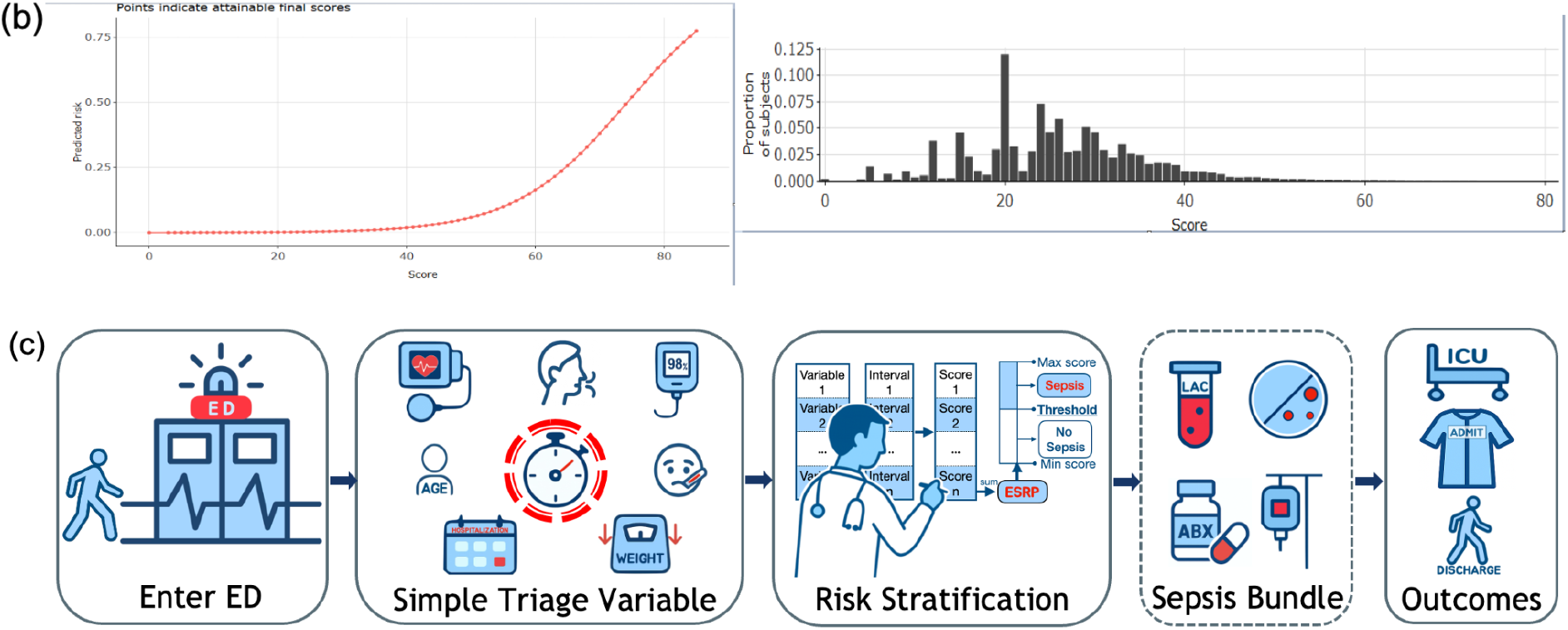
Performance and clinical workflow of the Emergency Sepsis Risk Prediction (ESRP) model. (a), Receiver operating characteristic curves of ESRP compared with benchmark models across 3 validation cohorts (University of Minnesota, Beth Israel Deaconess Medical Center, and Stanford). (b), Calibration curve (top) and score distribution histogram (bottom) of ESRP. (c), Conceptual workflow illustrating the use of ESRP in the emergency department, from patient presentation and triage variable collection to risk stratification, sepsis bundle initiation, and clinical outcomes. **Abbreviations:** AUC, indicates area under the curve; ESI, Emergency Severity Index; MEWS, Modified Early Warning Score; NEWS, National Early Warning Score; qSOFA, Quick Sequential Organ Failure Assessment; REMS, Rapid Emergency Medicine Score.

## Discussion

In this large, multicenter study of more than 2 million ED encounters, we developed and externally validated the ESRP score, a simple 10-variable tool derived from information routinely available from an initial ED evaluation. The ESRP demonstrated strong and consistent performance across diverse population and health systems, with AUROCs of 0.82 to 0.89, and outperformed commonly used bedside scores. Notably, its performance was comparable to more complex machine learning models while maintaining interpretability and ease of use. By relying only on information available at the time of triage and early ED evaluation, the ESRP provides an immediately actionable means of identifying patients at risk for sepsis, supporting earlier initiation of guideline-directed care in the ED.

### Strengths of the Study

This study has several notable strengths. First, the model was developed using a large-scale, real-world dataset of over one million ED encounters. Its generalizability was then rigorously validated on three separate validation cohorts from geographically diverse U.S. healthcare systems (Midwest, West Coast, and East Coast), which collectively included nearly one million additional patient encounters and exhibited significant differences in baseline characteristics and sepsis prevalence (from 0.7% to 2.5%). The ESRP was also validated across a wide spectrum of clinical environments, such as urban, suburban, and rural EDs, ensuring the model was developed and validated on heterogeneous patient populations. Second, the model achieved a strong balance between sensitivity (>74.3%) and specificity (>74.5%) across both internal and external validation cohorts. This indicates the ESRP score can effectively identify a high proportion of sepsis cases while achieving a reasonable balance between sensitivity and specificity, although the low PPV reflects a substantial proportion of false positives, which may limit its ability to minimize alarm fatigue and ensure clinical utility as a screening tool. Third, by leveraging the AutoScore algorithm^16^, we developed a model that is not only accurate but also highly interpretable and practical^23^, particularly when compared to less transparent models like RF and LR. The score is calculated using only 10 variables readily available during routine triage assessment, eliminating any delay for laboratory results. Its simple, point-based system allows for immediate risk calculation without requiring specialized expertise or imposing an additional workload on clinical staff. This design facilitates rapid, transparent, and actionable decision-making at the patient’s bedside.

### Interpretation of the ESRP Score Components

The clinical variables selected by the AutoScore algorithm for the ESRP score are all well-established risk factors for sepsis. Age was the most heavily weighted variable, with elderly patients receiving the highest score. This aligns with extensive literature demonstrating a higher incidence of sepsis in older adults^24^. The second-highest score was assigned to a chief complaint of fever or chills, which are cardinal signs of the systemic inflammatory response characteristic of sepsis^25,26^. Physiologic derangements at triage also contributed significantly to the score.

Hypotension is a hallmark of septic shock and tissue hypoperfusion and was rightly identified as a high-risk feature^27^. Similarly, tachycardia can be an early indicator of an infectious process or compensatory response^28^. Furthermore, tachypnea and hypoxemia are critical indicators of respiratory compromise, which may stem from a primary pulmonary infection that has triggered sepsis^29,30^, or acute lung injury resulting from sepsis. The inclusion of recent hospitalization is also a key risk factor, as associated hospital-acquired infections increase overall infection risk^31^. Beyond these foundational features, ESRP incorporates two comorbidities. Weight loss often serves as a proxy for underlying frailty, malnutrition, or malignancy—conditions known to predispose patients to severe infections, including sepsis^32^. Uncomplicated hypertension is also an independent risk factor^33^, which may predispose to sepsis by causing chronic endothelial dysfunction^34^. Based on the calibration analysis (eTable 5), the ESRP score can be stratified into three actionable risk groups. Patients with scores ≥67 (predicted risk ≥20%) represent a high-risk group warranting heightened index of suspicion for sepsis. Scores between 29 and 66 (0.5–20%) identify a medium-risk group for whom sepsis should be considered as a possible diagnosis, whereas scores <29 (<0.5%) define a low-risk group with excellent NPV, supporting consideration of non-sepsis diagnoses. This stratification framework facilitates risk-based management of suspected sepsis in clinical practice.

### Comparison with Existing Models and Scores

The ESRP score offers distinct advantages over existing clinical tools used for sepsis assessment. Although the full SOFA score is the cornerstone of the Sepsis-3 diagnostic criteria and has shown established prognostic value for ICU patients, in the ED setting the AUROC for SOFA score is around 0.700^35^. Its reliance on multiple laboratory results (e.g., PaO2/FiO2, platelet count, bilirubin, creatinine) that are not immediately available on patient presentation limit its use in rapid, frontline screening^25^. The qSOFA score was developed to fill this gap, but its simplicity is overshadowed by its widely criticized poor sensitivity^36^and the surviving sepsis guidelines now suggest against its use^37^. With a sensitivity reported to be as low as 16.3%, it leads to a high rate of missed sepsis cases, particularly in the early stages^38^. In contrast, our ESRP score is designed specifically for this ED triage window. It is not only laboratory-free, making it far more practical than SOFA for rapid screening, but it also achieves a higher predictive accuracy than qSOFA, addressing the key limitations of both tools. Other general risk scores like NEWS, MEWS, and REMS are designed to predict all-cause clinical deterioration or mortality^39^, but not sepsis specifically^40,41^. Consequently, they lack specificity. ESRP is purpose-built for sepsis, incorporating sepsis-specific features, which enhances its specificity.

When compared with other ML models, the ESRP score demonstrates a superior balance of performance and practicality. While some “black-box” models, such as those developed by Saqib et al., have achieved high AUROCs, their reliance on dozens of variables and lack of interpretability hinder clinical adoption^42^. More recent efforts have focused on interpretable models^43^, but they often face their own limitations. For example, a study by Liu et al. used SHAP for model explanation, yet their best-performing GB model required 62 input variables and achieved an AUROC of 0.83 in a large cohort of 189,617 patients with a sepsis prevalence of 5.95%^8^. A more direct comparison comes from the work of Jiang et al., who developed an interpretable “qSepsis” model using 12 variables without laboratory results. However, when tested on the same BIDMC external validation cohort used in our study, their model achieved an AUROC of only 0.766 (95% CI: 0.758–0.774)^44^. In stark contrast, our ESRP score relied on only 10 readily available variables and achieved a significantly higher AUROC of 0.838 (95% CI: 0.833–0.842) on this same dataset, highlighting its superior predictive accuracy and generalizability.

#### Limitations

We acknowledge several limitations. First, our study relies on administrative ICD codes and diagnosis description rather than the Sepsis-3 clinical criteria for outcome definition. Second, as a consequence of the low event rate in our development cohort, the AUROC may provide an optimistic view of performance. While the AUPRC is a more stringent metric in this context, its absolute value remains modest. Although our model’s AUPRC significantly exceeds the prevalence baseline, the value suggests that the rate of false positives remains a clinical consideration when implementing the score. Finally, this is a retrospective study, and its findings require prospective validation to confirm its clinical impact.

## Conclusions

We have developed and rigorously validated a simple, interpretable, and robust 10-variable scoring system, the ESRP score, for the early identification of sepsis in the ED. It balances high discriminative power, comparable to that of complex ML models, with the practicality required for frontline clinical use. By outperforming existing clinical scores, particularly in external validation, the ESRP score demonstrates its potential to become a valuable tool to improve the timely recognition of sepsis.

## Supporting information

eTable

## Data Availability

Data Availability Statement is in the manuscript

## Author contributions

YJ was responsible for data analysis, drafting of the manuscript, and critical revision. YW contributed to data preprocessing and management. MAP and DAW provided clinical expertise, interpretation of findings, and critical feedback on the manuscript. FX designed and supervised this study. All authors reviewed and approved the final version of the manuscript.

## Competing Interests

The authors declare no competing interests.

## Notes

### Competing Interest Statement

The authors have declared no competing interest.

### Funding Statement

This study did not receive any funding

### Author Declarations

This study was approved by the Institutional Review Board of the University of Minnesota (IRB #STUDY00023702).

## References

1. Yealy DM, Mohr NM, Shapiro NI, Venkatesh A, Jones AE, Self WH. Early care of adults with suspected sepsis in the emergency department and out-of-hospital environment: A consensus-based task force report. Ann Emerg Med. 2021;78(1):1–19.

2. Gaieski DF, Mikkelsen ME, Band RA, et al. Impact of time to antibiotics on survival in patients with severe sepsis or septic shock in whom early goal-directed therapy was initiated in the emergency department. Crit Care Med. 2010;38(4):1045–1053.

3. Wang W, Liu CF. Sepsis heterogeneity. World J Pediatr. 2023;19(10):919–927.

4. Lambden S, Laterre PF, Levy MM, Francois B. The SOFA score-development, utility and challenges of accurate assessment in clinical trials. Crit Care. 2019;23(1):374.

5. Askim Å, Moser F, Gustad LT, et al. Poor performance of quick-SOFA (qSOFA) score in predicting severe sepsis and mortality - a prospective study of patients admitted with infection to the emergency department. Scand J Trauma Resusc Emerg Med. 2017;25(1):56.

6. Williams B. The National Early Warning Score: from concept to NHS implementation. Clin Med. 2022;22(6):499–505.

7. Olsson T, Terent A, Lind L. Rapid Emergency Medicine score: a new prognostic tool for in-hospital mortality in nonsurgical emergency department patients. J Intern Med. 2004;255(5):579–587.

8. Liu Z, Shu W, Li T, Zhang X, Chong W. Interpretable machine learning for predicting sepsis risk in emergency triage patients. Sci Rep. 2025;15(1):887.

9. Fujimori R, Liu K, Soeno S, et al. Acceptance, barriers, and facilitators to implementing artificial intelligence-based decision support systems in emergency departments: Quantitative and qualitative evaluation. JMIR Form Res. 2022;6(6):e36501.

10. Xie F, Zhou J, Lee JW. Benchmarking emergency department prediction models with machine learning and public electronic health records. Sci Data. 2022;9. doi:10.1038/s41597-022-01782-9

11. Johnson AEW, Bulgarelli L, Shen L, et al. MIMIC-IV, a freely accessible electronic health record dataset. Sci Data. 2023;10(1):1.

12. Kansal A, Chen E, Jin T, Rajpurkar P, Kim D. Multimodal Clinical Monitoring in the Emergency Department (MC-MED). Published online 2025. doi:10.13026/JZ99-4J81

13. Kansal A, Chen E, Jin BT, Rajpurkar P, Kim DA. MC-MED, multimodal clinical monitoring in the emergency department. Sci Data. 2025;12(1):1094.

14. Haefeli M, Elfering A. Pain assessment. Eur Spine J. 2006;15 Suppl 1(S1):S17–S24.

15. Jain S, Margetis K, Iverson LM. Glasgow Coma Scale. In: StatPearls. StatPearls Publishing; 2025.

16. Xie F, Ning Y, Liu M, et al. A universal AutoScore framework to develop interpretable scoring systems for predicting common types of clinical outcomes. STAR Protoc. 2023;4(2):102302.

17. Xie F, Chakraborty B, Ong MEH, Goldstein BA, Liu N. AutoScore: A machine learning-based automatic clinical score generator and its application to mortality prediction using electronic health records. JMIR Med Inform. 2020;8(10):e21798.

18. Xie F, Ning Y, Yuan H, et al. AutoScore-Survival: Developing interpretable machine learning-based time-to-event scores with right-censored survival data. J Biomed Inform. 2022;125(103959):103959.

19. Modified Early Warning Score (MEWS). Accessed August 5, 2025. https://reference.medscape.com/calculator/461/modified-early-warning-score-mews

20. Xie F, Liu N, Yan L, et al. Development and validation of an interpretable machine learning scoring tool for estimating time to emergency readmissions. EClinicalMedicine. 2022;45(101315):101315.

21. Phungoen P, Khemtong S, Apiratwarakul K, Ienghong K, Kotruchin P. Emergency Severity Index as a predictor of in-hospital mortality in suspected sepsis patients in the emergency department. Am J Emerg Med. 2020;38(9):1854–1859.

22. Ozenne B, Subtil F, Maucort-Boulch D. The precision--recall curve overcame the optimism of the receiver operating characteristic curve in rare diseases. J Clin Epidemiol. 2015;68(8):855–859.

23. Xie F, Ong MEH, Liew JNMH, et al. Development and assessment of an interpretable machine learning triage tool for estimating mortality after emergency admissions. JAMA Netw Open. 2021;4(8):e2118467.

24. Clifford KM, Dy-Boarman EA, Haase KK, Maxvill K, Pass SE, Alvarez CA. Challenges with diagnosing and managing sepsis in older adults. Expert Rev Anti Infect Ther. 2016;14(2):231–241.

25. Singer M, Deutschman CS, Seymour CW, et al. The third international consensus definitions for sepsis and septic shock (sepsis-3). JAMA. 2016;315(8):801–810.

26. Tokuda Y, Miyasato H, Stein GH, Kishaba T. The degree of chills for risk of bacteremia in acute febrile illness. Am J Med. 2005;118(12):1417.

27. Maheshwari K, Nathanson BH, Munson SH, et al. The relationship between ICU hypotension and in-hospital mortality and morbidity in septic patients. Intensive Care Med. 2018;44(6):857–867.

28. Henning A, Krawiec C. Sinus tachycardia. In: StatPearls. StatPearls Publishing; 2025.

29. Wheeler AP, Bernard GR. Treating patients with severe sepsis. N Engl J Med. 1999;340(3):207–214.

30. Azoulay E, Russell L, Van de Louw A, et al. Diagnosis of severe respiratory infections in immunocompromised patients. Intensive Care Med. 2020;46(2):298–314.

31. Haque M, Sartelli M, McKimm J, Abu Bakar M. Health care-associated infections - an overview. Infect Drug Resist. 2018;11:2321–2333.

32. Lin W, Lin B, Chen J, et al. Impact of unintentional weight loss on 30-day mortality in intensive care unit sepsis patients: a retrospective cohort study. Sci Rep. 2024;14(1):31535.

33. Sun L, Zhang C, Song P, et al. Hypertension and 28-day mortality in sepsis patients: An observational and mendelian randomization study. Heart Lung. 2025;70:147–156.

34. Ahlberg CD, Wallam S, Tirba LA, Itumba SN, Gorman L, Galiatsatos P. Linking Sepsis with chronic arterial hypertension, diabetes mellitus, and socioeconomic factors in the United States: A scoping review. J Crit Care. 2023;77(154324):154324.

35. Innocenti F, Tozzi C, Donnini C, et al. SOFA score in septic patients: incremental prognostic value over age, comorbidities, and parameters of sepsis severity. Intern Emerg Med. 2018;13(3):405–412.

36. Kim KS, Suh GJ, Kim K, et al. Quick Sepsis-related Organ Failure Assessment score is not sensitive enough to predict 28-day mortality in emergency department patients with sepsis: a retrospective review. Clin Exp Emerg Med. 2019;6(1):77–83.

37. Evans L, Rhodes A, Alhazzani W, et al. Surviving sepsis campaign: International guidelines for management of sepsis and septic shock 2021. Crit Care Med. 2021;49(11):e1063–e1143.

38. Dorsett M, Kroll M, Smith CS, Asaro P, Liang SY, Moy HP. QSOFA has poor sensitivity for prehospital identification of severe sepsis and septic shock. Prehosp Emerg Care. 2017;21(4):489–497.

39. Yu JY, Xie F, Nan L, et al. An external validation study of the Score for Emergency Risk Prediction (SERP), an interpretable machine learning-based triage score for the emergency department. Sci Rep. 2022;12(1):17466.

40. Zhang K, Zhang X, Ding W, et al. National early warning score does not accurately predict mortality for patients with infection outside the intensive care unit: A systematic review and meta-analysis. Front Med (Lausanne). 2021;8:704358.

41. Adegbite BR, Edoa JR, Ndzebe Ndoumba WF, et al. A comparison of different scores for diagnosis and mortality prediction of adults with sepsis in Low-and-Middle-Income Countries: a systematic review and meta-analysis. EClinicalMedicine. 2021;42(101184):101184.

42. Saqib M, Sha Y, Wang MD. Early prediction of sepsis in EMR records using traditional ML techniques and deep learning LSTM networks. Annu Int Conf IEEE Eng Med Biol Soc. 2018;2018:4038–4041.

43. Yuan H, Yu K, Xie F, Liu M, Sun S. Automated machine learning with interpretation: A systematic review of methodologies and applications in healthcare. Medicine Advances. 2024;2(3):205–237.

44. Jiang S, Dai S, Li Y, et al. Development and validation of a screening tool for sepsis without laboratory results in the emergency department: a machine learning study. EClinicalMedicine. 2025;80(103048):103048.

