## Supplementary material for "Development and multicenter external validation of A Data-Driven Scoring System for Early and Rapid Identification of Sepsis in Emergency Departments": eTable

### Appendix

eMethod. Missing Data Assessment, Imputation, and AutoScore Preprocessing

eTable 1. Definitions of Study Variables by Category

eTable 2. List of Emergency Departments within the M Health Fairview System by Region

eTable 3. Baseline Characteristics of the UMN Internal Validation Cohort by Region

eTable 4. Performance of the ESRP Score in the UMN Internal Validation Cohort by Region

eTable 5. Performance Metrics of the ESRP Score at Different Predicted Risk Thresholds

eFigure 1. Calibration Curve for the ESRP Score after Platt Scaling

**eMethod. Missing Data Assessment, Imputation, and AutoScore Preprocessing**

We assessed the completeness of key variables prior to analysis. For instance, Systolic Blood Pressure (SBP), Diastolic Blood Pressure (DBP), and Mean Arterial Pressure (MAP) were each missing in just 1.47% of cases. Heart Rate was missing in 4.49% of records, while Temperature and Acuity Level had missing rates of 2.53% and 3.21%, respectively. Given these low rates, missing values for initial triage vital signs—SBP, DBP, temperature, heart rate, respiratory rate, and oxygen saturation (O2sat)—were imputed using the variable-specific median. Missing pain scores were imputed with 0, assuming the absence of documented pain, and missing Glasgow Coma Scale (GCS) scores were imputed with 15, assuming a normal neurological status if not otherwise recorded.

These preprocessing steps were conducted as preparation for AutoScore. After inputting variables with no missing values, AutoScore ranked them in descending order of importance using a machine learning model. We then selected the 10 variables with the strongest predictive impact, refined cutoffs for each variable based on clinical expertise or relevant literature, and assigned category-specific scores through the AutoScore framework. Model performance was evaluated using the AUROC, ultimately yielding the ESRP.

eTable 1: Definitions of Study Variables by Category

| Category | Variable Name | Description |
|---|---|---|
| Demographics | | |
| | Age | Age in year, specific to the index emergency admission |
| | Sex | Gender identity as identified in official patient identification documents |
| Medical History | | |
| | ED visits in past 30 days | Total number of ED visits within the past 30 days from the index emergency admission |
| | ED visits in past 90 days | Total number of ED visits within the past 90 days from the index emergency admission |
| | ED visits in past 1 year | Total number of ED visits within the past 1 year from the index emergency admission |
| | Hospitalizations in past 30 days | Total number of hospital admissions within the past 30 days from the index emergency admission (excluding observation status stays) |
| | Hospitalizations in past 90 days | Total number of hospital admissions within the past 90 days from the index emergency admission (excluding observation status stays) |
| | Hospitalizations in past 1 year | Total number of hospital admissions within the past 1 year from the index emergency admission (excluding observation status stays) |
| | ICU admissions in past 30 days | Total number of Intensive Care Unit (ICU) admissions within the past 30 days from the index emergency admission |
| | ICU admissions in past 90 | Total number of Intensive Care |

| | | |
|---|---|---|
| | days | Unit (ICU) admissions within the past 90 days from the index emergency admission |
| | ICU admissions in past 1 year | Total number of Intensive Care Unit (ICU) admissions within the past 1 year from the index emergency admission |
| Triage Vitals | | |
| | Temperature at triage | Patient’s body temperature recorded at triage, measured in Fahrenheit (˚F) or Celsius (˚C) |
| | Heart rate at triage | Heart rate recorded at triage, measured in beats per minute |
| | Respiratory rate at triage | Respiratory rate recorded at triage, measured in breaths per minute |
| | Oxygen saturation at triage | Oxygen saturation ($SpO_2$) recorded at triage, measured in percentage (%) |
| | Systolic BP at triage | Systolic blood pressure recorded at triage, measured in mmHg |
| | Diastolic BP at triage | Diastolic blood pressure recorded at triage, measured in mmHg |
| | Pain score at triage (0 - 10) | Self-reported pain score at triage on a scale from 0 (no pain) to 10 (worst pain imaginable) |
| | Triage acuity level (ESI) | Emergency Severity Index (ESI) triage acuity level assigned at triage |
| Chief Complaints | | |
| | Chief complaint: Chest pain | Patient’s primary reason for seeking care is pain in the chest area, as reported at triage. |
| | Chief complaint: Abdominal pain | Patient’s primary reason for seeking care is pain located in the abdominal region, as reported at triage. |
| | Chief complaint: Headache | Patient’s primary reason for seeking care is pain in the head |

| | | |
|---|---|---|
| | | region, as reported at triage. |
| | Chief complaint: Shortness of breath | Patient’s primary reason for seeking care is difficulty breathing or a sensation of not getting enough air, as reported at triage. |
| | Chief complaint: Back pain | Patient’s primary reason for seeking care is pain in the back region, as reported at triage. |
| | Chief complaint: Cough | Patient’s primary reason for seeking care is persistent or acute coughing, with or without sputum, as reported at triage. |
| | Chief complaint: Nausea/Vomiting | Patient’s primary reason for seeking care is feeling nauseous and/or actively vomiting, as reported at triage. |
| | Chief complaint: Fever/Chills | Patient’s primary reason for seeking care is having an elevated body temperature and/or experiencing chills, as reported at triage. |
| | Chief complaint: Syncope | Patient’s primary reason for seeking care is a transient loss of consciousness with spontaneous recovery, as reported at triage. |
| | Chief complaint: Dizziness | Patient’s primary reason for seeking care is a sensation of lightheadedness, unsteadiness, or vertigo, as reported at triage. |
| Comorbidities | | |
| | Myocardial infarction | ICD-9-CM: 410.x, 412.x; ICD-10-CM: I21.x, I22.x, I25.2 |
| | Congestive heart failure | ICD-9-CM: 398.91, 402.0x/402.1x/402.9x, 404.0x/404.1x/404.9x, 425.4–425.9, 428.x; ICD-10-CM: I09.9, I11.0, I13.2, I25.5, I42.0–I42.9, I43, I50.x, P29.0 |
| | Peripheral vascular disease | ICD-9-CM: 093.0, 437.3, 440.x, |

| | | |
|---|---|---|
| | | 441.x, 443.1–443.9, 447.1, 557.1, 557.9, V43.4; ICD-10-CM: I70.x, I71.x, I73.1, I73.8, I73.9, I77.1, I79.0, I79.2, K55.1, K55.8, K55.9, Z95.8, Z95.9 |
| | Cerebrovascular disease (stroke) | ICD-9-CM: 362.34, 430–438.x; ICD-10-CM: H34.0, G45.x, G46.x, I60–I69.x |
| | Dementia | ICD-9-CM: 290.x, 294.1, 331.2; ICD-10-CM: F00.x–F03.x, F05.1, G30.x, G31.1 |
| | Chronic pulmonary disease | ICD-9-CM: 490–505.x, 416.8–416.9, 506.4, 508.1, 508.8; ICD-10-CM: J40–J47, J60–J67, I27.8–I27.9, J68.4, J70.1, J70.3 |
| | Rheumatic disease | ICD-9-CM: 446.5, 710.0–710.4, 714.0–714.2, 714.8, 725; ICD-10-CM: M05.x, M06.x, M31.5, M32.x–M34.x, M35.1, M35.3, M36.0 |
| | Peptic ulcer disease | ICD-9-CM: 531.x–534.x; ICD-10-CM: K25.x–K28.x |
| | Mild liver disease | ICD-9-CM: 070.22, 070.23, 070.32, 070.33, 070.44, 070.54, 070.6, 070.9, 573.3, 573.4, 573.8, 573.9, V42.7, 570, 571.x; ICD-10-CM: K70.0–K70.3, K70.9, K71.3–K71.5, K71.7, K76.0, K76.2–K76.4, K76.8–K76.9, Z94.4, B18.x, K73.x, K74.x |
| | Diabetes without complications | ICD-9-CM: 250.0–250.3, 250.8–250.9; ICD-10-CM: E10.0–E10.1, E10.6, E10.8–E10.9; E11.0–E11.1, E11.6, E11.8–E11.9; E12.0–E12.1, E12.6, E12.8–E12.9; E13.0–E13.1, E13.6, E13.8–E13.9; E14.0–E14.1, E14.6, E14.8–E14.9 |
| | Diabetes with complications | ICD-9-CM: 250.4–250.7; ICD-10-CM: E10.2–E10.5, E10.7; E11.2– |

| | | |
|---|---|---|
| | | E11.5, E11.7; E12.2–E12.5, E12.7; E13.2–E13.5, E13.7; E14.2–E14.5, E14.7 |
| | Hemiplegia or paraplegia | ICD-9-CM: 334.0–334.6, 334.9, 334.1, 342.x, 343.x; ICD-10-CM: G04.1, G11.4, G80.1–G80.2, G81.x, G82.x, G83.0–G83.4, G83.9 |
| | Renal disease | ICD-9-CM: 403.01/403.11/403.91, 404.02/404.03/404.12/404.13/404.92/404.93, 582.x, 583.0–583.7, 585, 586, 588.0, V42.0, V45.1, V56.x; ICD-10-CM: I12.0, I13.1, N03.2–N03.7, N05.2–N05.7, N18.x, N19, N25.0, Z49.0–Z49.2, Z94.0, Z99.2 |
| | Solid tumor without metastasis | ICD-9-CM: 140–195.x, 238.6; ICD-10-CM: C00–C26, C30–C41, C43, C45–C49, C50–C58, C60–C76, C97 |
| | Moderate or severe liver disease | ICD-9-CM: 456.0–456.2, 572.2–572.8; ICD-10-CM: I85.0, I85.9, I86.4, I98.2, K70.4, K71.1, K72.1, K72.9, K76.5–K76.7 |
| | Metastatic solid tumor | ICD-9-CM: 196–199.x; ICD-10-CM: C77–C80 |
| | HIV/AIDS | ICD-9-CM: 042–044; ICD-10-CM: B20–B24 |
| | Cardiac arrhythmias | ICD-9-CM: 426.0, 426.10, 426.12–426.13, 426.7, 426.9, 427.0–427.4, 427.6–427.9, 785.0, 996.01, 996.04, V45.0, V53.3; ICD-10-CM: I44.1–I44.3, I45.6, I45.9, I47.x–I49.x, R00.0–R00.1, R00.8, T82.1, Z45.0, Z95.0 |
| | Valvular disease | ICD-9-CM: 093.2, 394–397.x, 424.x, 746.3–746.6, V42.2, V43.3; ICD-10-CM: A52.0, I05– |

| | | |
|---|---|---|
| | | I08, I09.1, I09.8, I34–I39, Q23.0–Q23.3, Z95.2–Z95.4 |
| | Pulmonary circulation disorders | ICD-9-CM: 415.0–415.1, 416.x, 417.0, 417.8–417.9; ICD-10-CM: I26.x, I27.x, I28.0, I28.8–I28.9 |
| | Hypertension, uncomplicated | ICD-9-CM: 401.x; ICD-10-CM: I10 |
| | Hypertension, complicated | ICD-9-CM: 402.x, 403.x, 404.x, 405.x; ICD-10-CM: I11, I12, I13, I15 |
| | Other neurological disorders | ICD-9-CM: 331.9, 332.0–332.1, 333.4–333.5, 333.92, 334.x, 335.x, 336.2, 340.x, 341.x, 345.x, 348.1, 348.3, 780.3, 784.3; ICD-10-CM: G10–G13, G20–G22, G32, G35–G37, G40–G41, R56, G25.4–G25.5, G31.2, G31.8–G31.9, G93.1, G93.4, R47.0 |
| | Hypothyroidism | ICD-9-CM: 240.9, 243, 244.x, 246.1, 246.8; ICD-10-CM: E00–E03, E89.0 |
| | Lymphoma | ICD-9-CM: 200–202.x, 203.0, 238.6; ICD-10-CM: C81–C85, C88, C90.0, C90.2, C96 |
| | Coagulopathy | ICD-9-CM: 286.x, 287.1, 287.3–287.5; ICD-10-CM: D65–D68, D69.1, D69.3–D69.6 |
| | Obesity | ICD-9-CM: 278.0; ICD-10-CM: E66.x |
| | Weight loss | ICD-9-CM: 783.2, 799.4, 260–263.x; ICD-10-CM: R63.4, R64, E40–E46 |
| | Fluid and electrolyte disorders | ICD-9-CM: 253.6, 276.x; ICD-10-CM: E86–E87 |
| | Blood loss anemia | ICD-9-CM: 280.0; ICD-10-CM: D50.0 |
| | Deficiency anemia | ICD-9-CM: 280.1–280.9, 281.x; ICD-10-CM: D50.8–D50.9, D51–D53 |
| | Alcohol abuse | ICD-9-CM: 291.1, 291.3, 291.5– |

| | | |
|---|---|---|
| | | 291.9, 303.0, 303.9, 305.0, 357.5, 425.5, 535.3, 571.0–571.3, V11.3, 980.x; ICD-10-CM: F10.x, E52, G62.1, I42.6, K29.2, K70.0, K70.3, K70.9, T51.x, Z50.2, Z71.4, Z72.1 |
| | Drug abuse | ICD-9-CM: 292.x, 304.x, 305.2–305.9, V65.42; ICD-10-CM: F11–F16, F18–F19, Z71.5, Z72.2 |
| | Psychoses | ICD-9-CM: 293.8, 295.x, 297.x, 298.x, 296.04, 296.14, 296.44, 296.54; ICD-10-CM: F20–F25, F28–F29, F30.2, F31.2, F31.5 |
| | Depression | ICD-9-CM: 296.2, 296.3, 296.5, 300.4, 311, 309.x; ICD-10-CM: F32.x, F33.x, F34.1, F41.2, F43.2, F31.3–F31.5 |
| Outcome | | |
| | Sepsis | ICD-9-CM: 99591, 99592, 67024; ICD-10-CM: A419, R6520, A4189, R6521, O0387, T8144XA, O0337, O0487, A4150, A410, A411, A412, A403, A414, A4151, A4152 |

eTable 2. List of Emergency Departments within the M Health Fairview System by Region

| | Region | City | Hospital Name |
|---|---|---|---|
| 1 | Urban | Minneapolis | Emergency Department - M Health Fairview Masonic Children’s Hospital |
| 2 | Urban | Minneapolis | Emergency Department - M Health Fairview University of Minnesota Medical Center - East Bank |
| 3 | Urban | Minneapolis | Emergency Department - M Health Fairview University of Minnesota Medical Center - West Bank |
| 4 | Suburban | Burnsville | Emergency Department - M Health Fairview Ridges Hospital |
| 5 | Suburban | Edina | Emergency Department - M Health Fairview Southdale Hospital |
| 6 | Suburban | Maplewood | Emergency Department - M Health Fairview St. John's Hospital |
| 7 | Suburban | Woodbury | Emergency Department - M Health Fairview Woodwinds Hospital |
| 8 | Rural | Grand Rapids | Grand Itasca Clinic and Hospital |
| 9 | Rural | Hibbing | Fairview Range Medieal Center |
| 10 | Rural | Princeton | Emergency Department - M Health Fairview Northland Medical Center |
| 11 | Rural | Wyoming | Emergency Department - M Health Fairview Lakes Medical Center |

eTable 3: Baseline Characteristics of the UMN Internal Validation Cohort by Region

| **Cohort** | Urban | Suburban | Rural |
|---|---|---|---|
| Sample Size (N) | 55,705 | 200,000 | 93,169 |
| Age, Mean (SD) (years) | 45.37 (19.26) | 53.55 (21.37) | 52.80 (20.69) |
| Gender, Female (%) | 30,765 (55.2) | 117127 (57.2) | 51,595 (55.4) |
| Gender, Male (%) | 24,940 (44.8) | 87504 (42.8) | 41,574 (44.6) |
| ESI, Level 1 (%) | 230 (0.4) | 1,567 (0.8) | 468 (0.5) |
| ESI, Level 2 (%) | 11,827 (21.2) | 53,922 (26.4) | 14,002 (15.0) |
| ESI, Level 3 (%) | 35,533 (63.8) | 116,952 (57.2) | 64,277 (69.0) |
| ESI, Level 4 (%) | 7,336 (13.2) | 30,632 (15.0) | 13,356 (14.3) |
| ESI, Level 5 (%) | 779 (1.4) | 1,558 (0.8) | 1,066 (1.1) |
| Systolic BP at Triage, Mean (SD) (mmHg) | 126.66 (18.67) | 133.47 (21.08) | 134.37 (20.36) |
| Diastolic BP at Triage, Mean (SD) (mmHg) | 77.88 (12.62) | 77.44 (13.76) | 80.18 (13.89) |
| Temperature at Triage, Mean (SD) (°C) | 36.73 (0.39) | 36.69 (0.43) | 36.72 (0.46) |
| Heart Rate at Triage, Mean (SD) (bpm) | 81.68 (15.37) | 79.35 (15.07) | 81.03 (15.47) |
| Oxygen Saturation at Triage, Mean (SD) (%) | 97.68 (2.95) | 97.14 (2.86) | 96.92 (2.53) |
| Respiratory Rate at Triage, Mean (SD) (breaths/min) | 17.48 (2.76) | 18.39 (3.08) | 18.26 (3.19) |
| Pain Score at Triage, Mean (SD) (0–10 scale) | 0.00 (0.00) | 0.00 (0.00) | 0.00 (0.00) |
| Sepsis, Positive (%) | 379 (0.7) | 1,866 (0.9) | 724 (0.8) |

eTable 4: Performance of the ESRP Score in the UMN Internal Validation Cohort by Region

| UMN data - internal validation cohort | | | | | | |
|---|---|---|---|---|---|---|
| **Model** | **Threshold** | **AUROC** | **AUPRC** | **Sensitivity** | **Specificity** | **Variables (N)** |
| ESRP Score - Urban | 36 | 0.805 (0.778–0.82) | 0.05 (0.045–0.07) | 0.731 (0.682–0.775) | 0.768 (0.734–0.77) | 10 |
| ESRP Score - Suburban | 36 | 0.815 (0.807–0.823) | 0.057 (0.053–0.062) | 0.735 (0.728–0.792) | 0.735 (0.694–0.737) | 10 |
| ESRP Score - Rural | 37 | 0.837 (0.829–0.851) | 0.055 (0.05–0.071) | 0.733 (0.72–0.791) | 0.799 (0.75–0.801) | 10 |

eTable 5: Performance Metrics of the ESRP Score at Different Predicted Risk Thresholds

| Predicted Risk [>=] | Score cut-off [>=] | Percentage of patients (%) | Accuracy (95% CI) | Sensitivity (95% CI) | Specificity (95% CI) | PPV (95% CI) | NPV (95% CI) |
|---|---|---|---|---|---|---|---|
| 0.10% | 14 | 92 | 8.7% (8.6-8.8%) | 99.4% (99.1-99.7%) | 7.9% (7.8-8%) | 0.9% (0.9-0.9%) | 99.9% (99.9-100%) |
| 0.50% | 29 | 43 | 57.5% (57.4-57.7%) | 87.3% (86-88.4%) | 57.3% (57.1-57.4%) | 1.7% (1.7-1.7%) | 99.8% (99.8-99.8%) |
| 1% | 36 | 20 | 80.2% (80-80.3%) | 66.5% (64.8-68.2%) | 80.3% (80.2-80.4%) | 2.8% (2.7-2.8%) | 99.6% (99.6-99.7%) |
| 10% | 59 | 1 | 98.7% (98.7-98.7%) | 8.7% (7.7-9.8%) | 99.5% (99.5-99.5%) | 12.5% (11.1-13.9%) | 99.2% (99.2-99.2%) |
| 20% | 67 | 0 | 99.1% (99.1-99.1%) | 2.2% (1.7-2.7%) | 99.9% (99.9-99.9%) | 15% (11.8-18.6%) | 99.2% (99.2-99.2%) |

eFigure 1: Calibration Curve for the ESRP Score after Platt Scaling

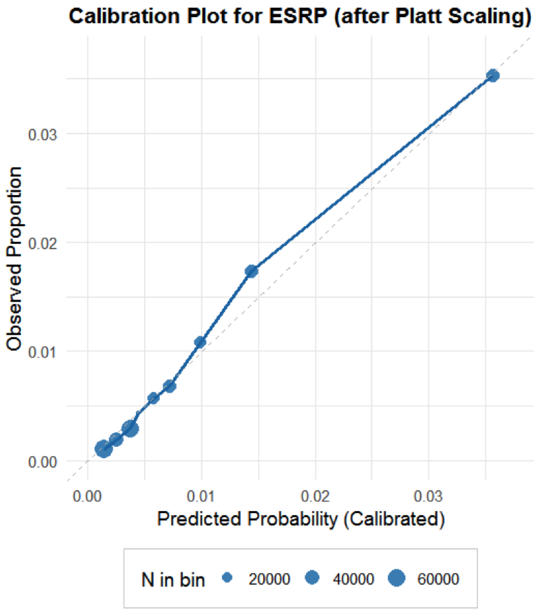
